# Initial Results with an Absorbable Urologic Scaffold to Mitigate Early Urinary Incontinence Following Radical Prostatectomy: The ARID Study

**DOI:** 10.1101/2025.06.29.25330348

**Authors:** Jeffrey C. Gahan, Gustavo Espino, Marcos Young, Elías Bodden, Michael N. Ferrandino

**Author notes:** Corresponding Author: Jeffrey C. Gahan, MD Department of Urology Duke Heath, 3404 Wake Forest Road Raleigh, NC 27609-7340. Clinical Trial Registration Number: NCT06275945 Registration Date: February 23, 2024. Initial results from this study were presented at the American Urological Association 2025 Annual Meeting, April 26-29, 2025.

## Abstract

**Purpose:** Stress urinary incontinence (SUI) is a frequent adverse effect following robot-assisted radical prostatectomy (RARP) for prostate cancer. Causes include urethra shortening, bladder neck widening and diminished urethral support. To address these, an implantable, absorbable urologic scaffold has been developed to elongate the urethra and to provide radial support to the bladder neck and urethral stump, at the time of RARP.

**Methods:** Prospective, non-randomized, single-arm study with the urologic scaffold placed at the time of RARP anastomosis. Endpoints include continence rate at differing timepoints up to 6 months and adverse event rate. Continence was rigorously defined as return to pre-surgery pad weight, inclusive of measurement error, using 24-hour pad weight testing.

**Results:** Twenty-four subjects with a mean age of 62.9±7.4 years and BMI of 27.1±3.5 were enrolled. Half of the subjects were continent upon catheter removal with 52.2% and 76.2% being continent at 6 weeks and 6 months based on 24-hour pad weight testing. Sub-optimal device placement was determined by video review in some subjects. A sub-analysis was performed which showed 80.0% and 92.9% continence rates at 6 weeks and 6 months for subjects with correctly placed devices compared to 0% and 42.9% when sub-optimally placed. No device-related adverse events were reported.

**Conclusion:** Early experience with an absorbable urologic scaffold demonstrates the device is safe and reduces SUI following RARP with proper placement associated with improved outcomes Longer term results from this study and an ongoing randomized controlled trial will further define the device’s role towards reducing SUI after RARP.

## Introduction

Stress urinary incontinence is a frequent adverse effect following radical prostatectomy (RP) for prostate cancer [1]. Reports indicate between 66% to 80% of men experience continued incontinence at 3 months post RP [2,3] and 5% to 20% will go on to develop long-term incontinence [1,4,5] An increase in the number of risk factors for incontinence prior to undergoing RP is associated with a higher likelihood that a patient will remain incontinent at 2 years [6]. A recent study focusing on the long-term adverse effects and complications after prostate cancer treatment reported a 5 times increased risk of urinary incontinence during the 12 years following RP compared to patient undergoing radiotherapy [7].

Stress urinary incontinence is predominantly related to inherent limitations of the prostatectomy technique. Surgical removal of the prostate leads to a shortened urethra, widened bladder neck, and diminished urethral support. This results in increased demand on the urinary sphincter to maintain urinary continence. While multiple risk factors have been shown to be associated with prolonged stress incontinence following RP, many are not modifiable including patient age, prostate volume, urethral length, and importantly, surgeon experience [7]. As a result, newer options designed to prevent stress urinary continence during the perioperative period are needed.

An absorbable scaffold (Voro Urologic Scaffold, Levee Medical, Durham, NC) has been developed to provide radial support to the bladder neck and urethral stump at the anastomosis to effectively elongate the urethra proximal to the urinary sphincter [8]. This, in turn, reduces the stresses on the urinary sphincter to help quickly restore urinary continence following surgery. The scaffold is composed of polydioxanone (PDO) monofilament, similar in form to PDO sutures and poly L-lactide/caprolactone (PLC) to form an atraumatic hem used to terminate the distal braided monofilament (Fig 1). The device is designed to be efficiently placed at the time of prostatectomy utilizing standard equipment and surgical techniques and is intended to reduce the occurrence and severity of post-prostatectomy stress urinary incontinence. The device is fully absorbable and serves as a scaffold for organized tissue healing that helps maintain the favorable bladder neck geometry.

**Fig. 1.**
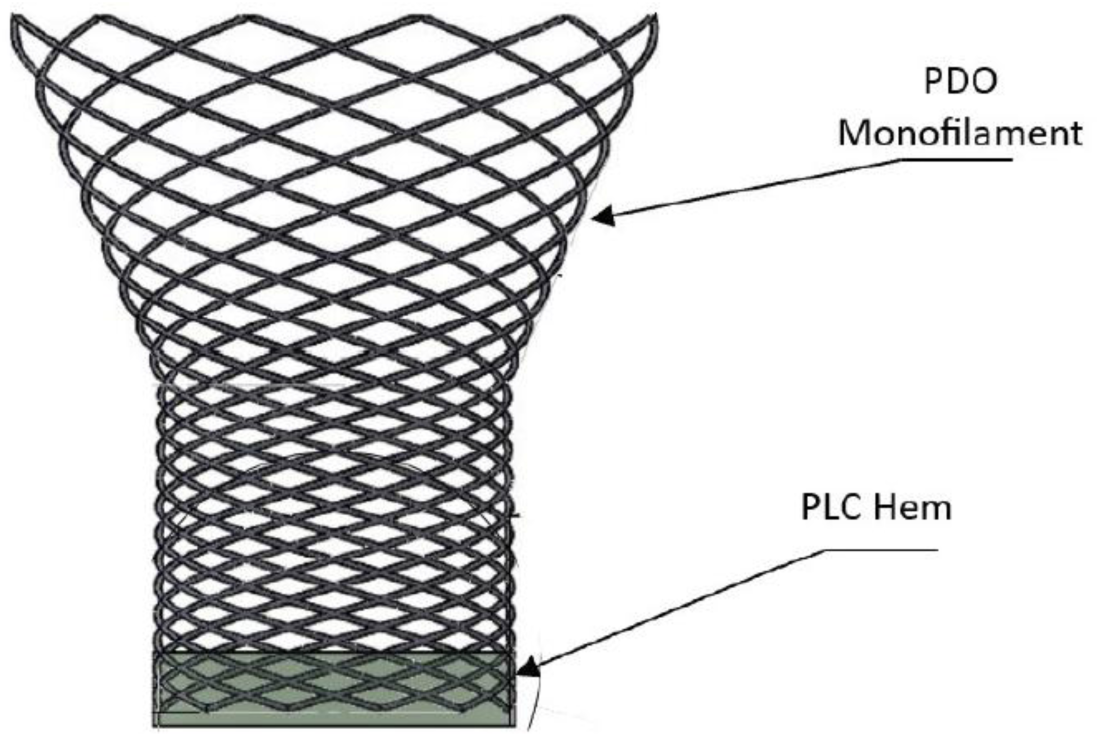
The Voro Urologic Scaffold

Presented here are the interim 6 months findings from the ARID study (NCT06275945), a prospective trial evaluating the safety and performance of a novel absorbable urologic scaffold designed to prevent stress urinary incontinence in patients undergoing RARP for the treatment of prostate cancer.

## Materials and Methods

### Study Design

The ARID study is a prospective, single-center, early feasibility clinical trial of an implantable absorbable urologic scaffold in men with prostate cancer undergoing RARP. The objective of this study is to evaluate the safety and performance of the urologic scaffold as a prophylactic treatment for post-prostatectomy stress urinary incontinence. The study was approved by the Comité de Bioética en Investigación at Pacífica Salud Hospital Punta Pacífica and all subjects signed a written informed consent to participate in the study and undergo surgery with the placement of the absorbable scaffold.

Patient demographics, disease information, procedural data, and post-operative outcomes and safety data were collected prospectively.

The study enrolled adult males 45 to 70 years of age diagnosed with Gleason Grade Group 3 or lower prostate cancer, prostate size less than 80 grams who were scheduled for radical prostatectomy. Subjects were excluded if they had a high suspicions of extra prostatic extension, a history of urinary incontinence, taking medications to treat overactive bladder, a post void residual greater than either 200 ml or less than 25% voiding efficiency, the presence of urethral stricture or bladder neck contracture, current or chronic urinary tract infection, prior urologic surgery or minimally invasive procedures, prior pelvic radiation or anticipated need for radiation after radical prostatectomy, the presence of stones in the bladder, a history of neurogenic bladder, sphincter abnormalities, or poor detrusor muscle function, a body mass index greater than 35, a history of other cancer, excluding prostate cancer, not considered in complete remission, diagnosed or suspected primary neurologic conditions known to affect bladder function, sphincter function or poor detrusor muscle function, a history of clinically significant congestive heart failure, insulin-dependent diabetes mellitus or uncontrolled diabetes, intravesical prostatic protrusion greater than 5mm, a history of immunosuppressive conditions or any other significant medical history or other condition which makes the subject unsuitable for the study per investigator discretion.

### Endpoints

The primary efficacy endpoint for this interim analysis of data from ARID study was the change in average 24-hour pad weight over 6 months. Subjects are reported as being continent if their post-surgery pad weight measurements were less than or equal to their pre-surgery values inclusive of the measurement error. The measurement error was determined to be three standard deviations of the pre-surgery 24-hour pad weight test calculated based on the results reported.

The secondary study endpoint related to change in pad weight during a 1-hour provocative pad weight test was also analyzed. The 1-hour pad test followed the standardized testing protocol defined by the International Continence Society [9]. The primary safety endpoint was the rate of serious adverse events (SAEs) and adverse events (AEs) in the analysis population based on Common Terminology Criteria for Adverse Events (CTCAE v5.0) [10].

### Surgical Technique

Following excision of the prostate, the urologic scaffold was compressed and introduced into the surgical field through a trocar using standard laparoscopic graspers. The smaller diameter end of the scaffold was oriented facing the urethral stump, and while compressed, was placed over the stump at the pelvic floor (Fig. 2).

**Fig. 2.**
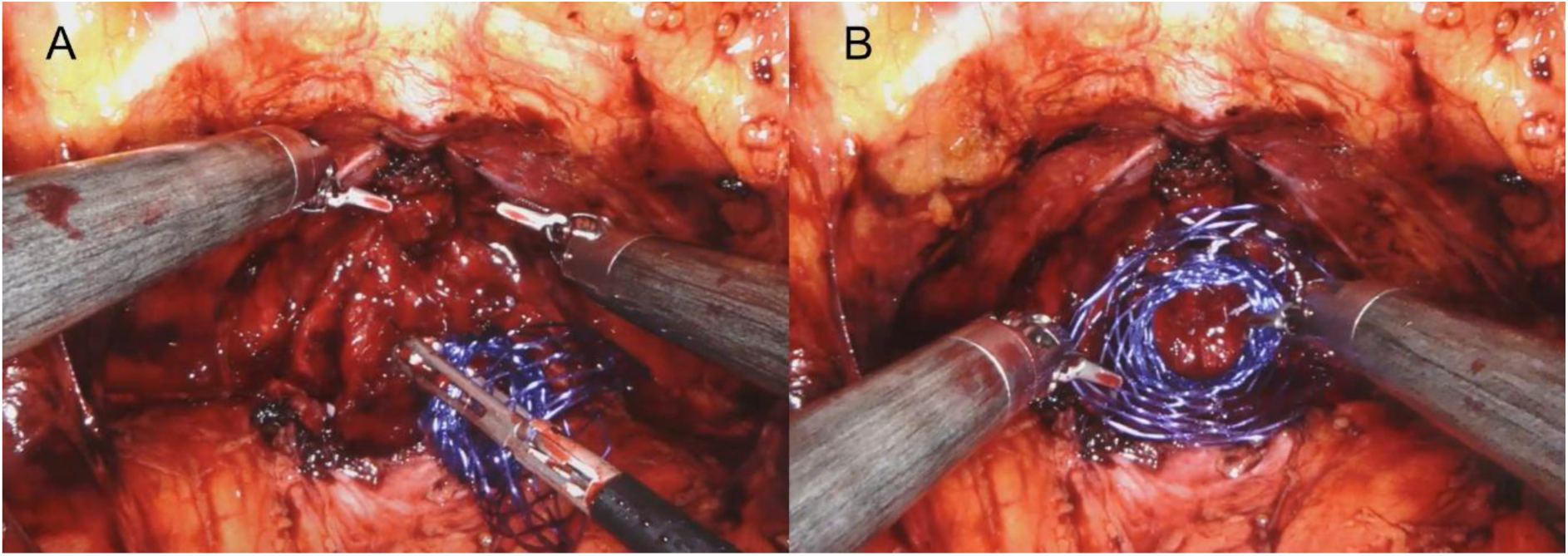
Prior to performing the anastomosis, the urologic scaffold (purple) is inserted in compressed state **(A)**. Following placement over the ureteral stump, the urethra is shown projecting through the central lumen of the scaffold prior to anastomosis **(B)**. The anastomosis is subsequently performed through the device.

After correct positioning, the anastomosis between the urethra and the bladder neck was completed through the center of the scaffold using barbed sutures. Following a negative leak test, the scaffold was expanded and the base secured to the urethra using two absorbable sutures positioned opposite of each other. The scaffold was then expanded towards the bladder neck using gentle lateral compression along each side of the device. Once completely expanded and sized to ensure urethral support, two sutures were placed near the midpoint of the device to prevent it from sliding distally. The widest portion of the scaffold was then secured to the bladder neck using two anchoring sutures, ensuring the suture passed through muscular tissues for proper anchoring (Figs. 3 and 4). A final leak test was to ensure the desired bladder neck geometry had been achieved. Incisions were closed according to the standard of care for radical prostatectomy procedures.

**Fig. 3.**
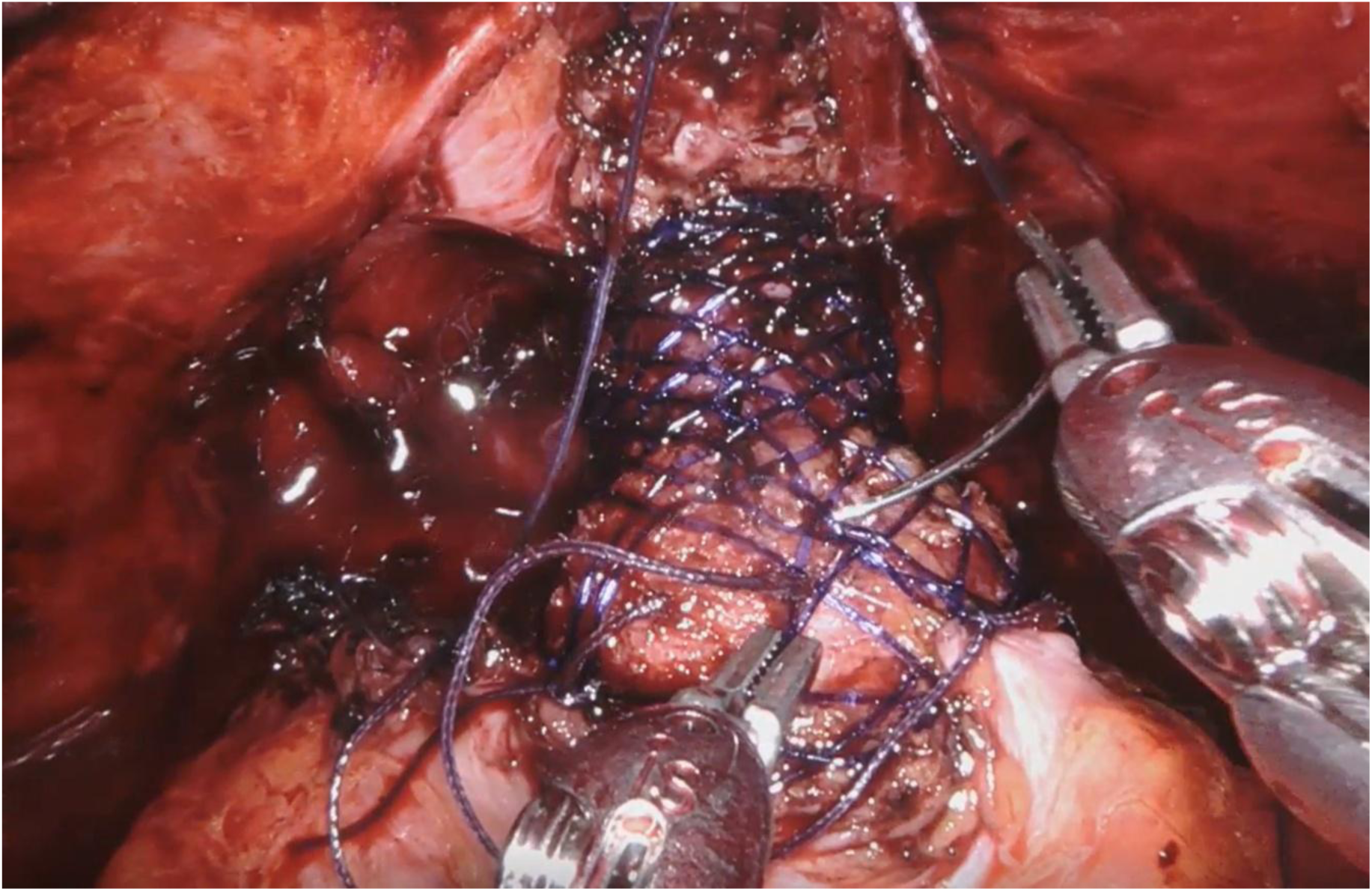
Suture placement to secure urologic scaffold in place following creation of the anastomosis.

**Fig. 4.**
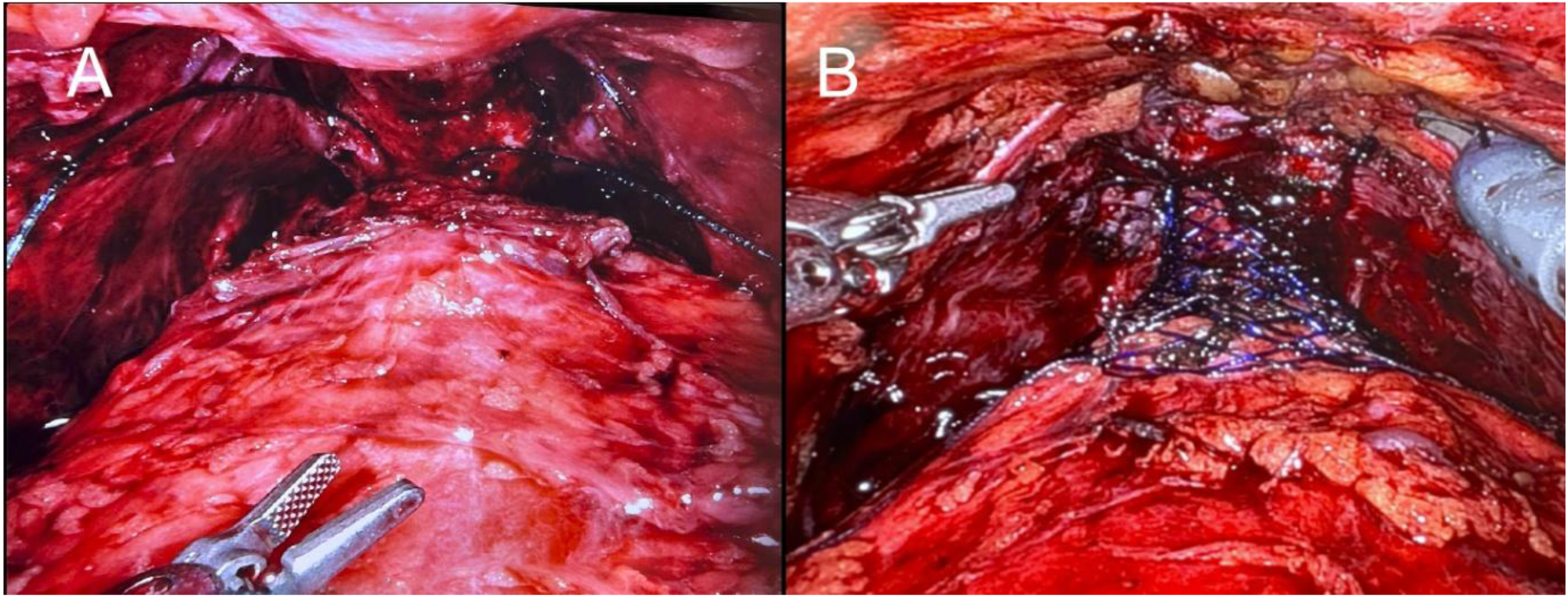
Image of anastomosed bladder **(A)** without and **(B)** with Voro Urologic Scaffold

### Statistical Analysis

There are no statistically powered endpoints for the ARID study and therefore a sample size determination was not performed. Descriptive methods are used to summarize continuous variables (e.g., mean, standard deviation, minimum, and maximum), and frequency tables or proportions are used to summarize categorical variables. A 95% confidence interval (CI) was constructed when deemed necessary.

## Results

Between April 2023 and April 2024 a total of 33 patients were pre-screened for the study and 29 patients signed informed consent forms. Four patients were screen failures based on the study’s inclusion and exclusion criteria and one patient was withdrawn from the study during the procedure prior to device placement due to the bilateral ureteral orifices being very close to the bladder neck. The first 4 subjects who were implanted with urologic scaffold are excluded from the present analysis since they received an initial version of the device which was limited in its ability to fully extend and support the bladder neck due to its large size. The remaining 24 subjects were implanted with a smaller version of the device and included in the present analysis. Details on study enrollment and exclusions are provided in the CONSORT diagram (Fig. 5).

**Fig. 5.**
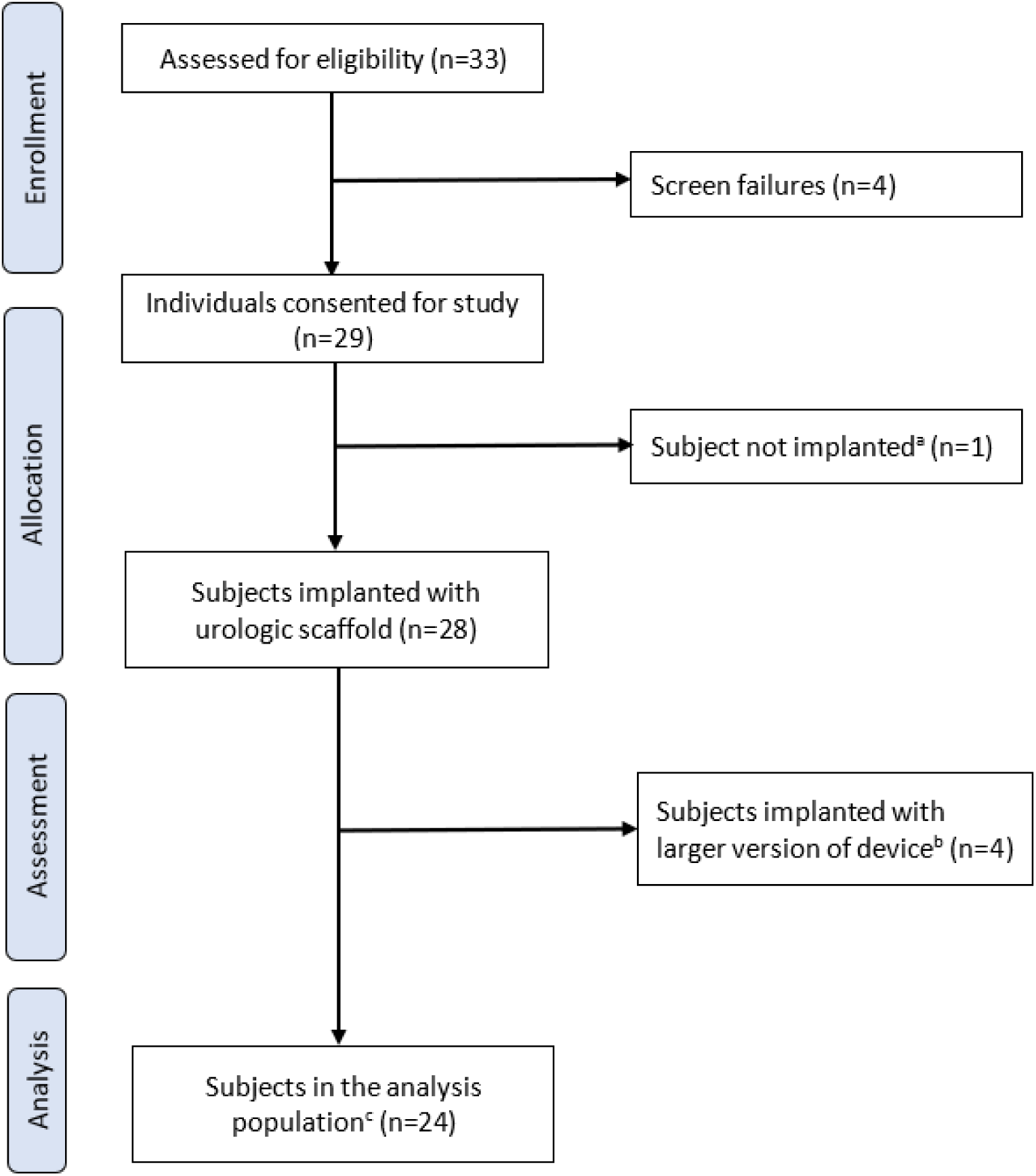
CONSORT flow diagram for ARID Study

Baseline subject demographics and characteristics are listed in Table 1. The mean age was 62.9 ± 7.4 years and mean BMI was 27.1 ± 3.5. Eight of 24 subjects (33.3%) were Gleason Grade Group 1, 10 (41.7%) were Grade Group 2, and 6 (25.0%) were Grade Group 3.

**Table 1.** Patient demographics and clinical characteristics.

|  |  |
| --- | --- |
| Number of subjects enrolled | 24 |
| Age, years ± SD (range) | $62.9 \pm 7.4$ (45, 70) |
| Male gender, number (%) | 29/29 (100%) |
| BMI | $27.1 \pm 3.5$ (21.2, 33.4) |
| Prostate size, mL ± SD (range) | $28.6 \pm 14.2$ (14.2, 73.4) |
| Serum Creatinine, mg/dL ± SD (range) | $1.0 \pm 0.2$ (0.7, 1.5) |
| Hemoglobin, g/dL ± SD (range) | $15.0 \pm 0.8$ (13.4, 16.2) |
| Hematocrit, % ± SD (range) | $44.0 \pm 2.5$ (38.8, 48.2) |
| Gleason Grade Group, number (%) |  |
| Grade 1 | 8 (33.3%) |
| Grade 2 | 10 (41.7%) |
| Grade 3 | 6 (25.0%) |

### Primary Endpoint: 24-hour Pad Weight Test

The ARID Study assessed the degree of each subject’s incontinence by comparing pad weights before and after prostatectomy. Pad weight measurements were obtained prior to each subject undergoing the prostatectomy to establish a baseline and at each specified time point following prostatectomy (Supplementary Table 1). Subjects were classified as being continent if their post-surgery pad weight measurements were less than or equal to their pre-surgery values inclusive of the measurement error of the pad weight test. Since pad weight tests are known to have high within-patient variability [11], the ARID study accounted for this variability by defining subjects as being continent if their change in pad weight from baseline was within 34.8 grams. This value is equal to three standard deviations of the pre-surgery pad weight values of the analysis population. This approach enabled a strict and objective definition of urinary continence.

Table 2 summarizes the percentage of subjects achieving continence at each timepoint of the study. 24-hour pad weight data at catheter removal was not available for 5 subjects as this data point was not being collected in an earlier version of the protocol. Pad weight data was also not available at the time of this report for 3 subjects for the 3 and 6 month follow-up visits. Data for the 4 to 6 week visit for one subject was excluded from the analysis because they presented with a meatal stenosis which was deemed to be a confounding variable. Overall, 57.1% of subjects were reported as being continent at 3 months and 76.2% at 6 months.

**Table 2.** Continence rates based on 24-hour pad weight testing, by study visit, n (%)

|  | Catheter removal<sup>a</sup> | 4-6 weeks | 3 months<sup>c</sup> | 6 months<sup>c</sup> |
| --- | --- | --- | --- | --- |
| All subjects | 9/19 (47.4%) | 12/23 (52.2%)<sup>b</sup> | 12/21 (57.1%) | 16/21 (72.2%) |
| Subjects with full device extension | 9/12 (75.0%) | 12/15 (80.0%)<sup>b</sup> | 12/14 (85.7%) | 13/14 (92.9%) |
| Subjects with minimal device extension | 0/7 (0.0%) | 0/8 (0.0%) | 0/7 (0.0%) | 3/7 (42.9%) |

Values reported as number of evaluable subjects at each time point (%)
<sup>a</sup> Visit not required for 5 subjects initially enrolled in a previous version of the study protocol
<sup>b</sup> One subject not included in the analysis because they presented with a meatal stenosis at the penis tip due to catheter placement.
<sup>c</sup> Data not yet available for 3 subjects at time of cutoff date for present report.

A sub-analysis of the 24-hour pad weight data was conducted after video review of the scaffold placement position showed suboptimal adherence to the standardized deployment technique for some subjects. The sub-analysis demonstrated a discernable difference in the percentage of subjects achieving continence at each time point depending on whether the urologic scaffold was fully extended or not (Table 2). Subjects who had fully extended devices experienced a 75.0%, 80.0%, 85.7% and a 92.9% continence rate at catheter removal and the 4 to 6 week, 3 month and 6 month visits, respectively. This compares to none of the subjects who had minimally extended devices achieving continence during the first 3 follow-up visits and only 42.9% achieving continence at 6 months.

### 1-hour Pad Weight Test

Similar to the analysis used for the 24-hour pad weight test, the difference in measured 1-hour pad weight between baseline and post-prostatectomy was used to determine if the subject returned to continence (Supplementary Table 2). To account for measurement error, which is smaller with the 1-hour compared to the 24-hour pad weight test, a subject was considered to have returned to baseline continence if there was less than 5 grams of weight gain versus basis for the 1-hour test.

Supplementary Table 2 summarizes the percentage of subjects achieving continence at each timepoint of the study based on 1-hour pad weight testing. Pad weight data was not available at the time of this report for 3 subjects for the 3 and 6 month follow-up visits. Data for the 4 to 6 week visit for one subject was excluded from the analysis because they presented with a meatal stenosis which was deemed to be a confounding variable. Overall, approximately 60% of subjects were reported as being continent during the 4 to 6 week, 3 month and 6 month follow-up visits (Table 3). Similar to the above, a separate sub-analysis was performed to compare results associated with the 1-hour pad weight test for subjects who had fully extended devices versus those with limited device extension. The sub-group analysis also found a positive relationship between device extension and patient continence outcomes with 87.5% of subjects with full device extension being continent at 6 weeks versus none (0.0%) of subjects who had minimally device extension (Table 3). This difference was also observed at both 3 and 6 months with a 78.6% continence rate at both time points for subjects with fully extended devices compared to 14.3% of subjects who had suboptimal device placement. All subjects (100%) who had optimally placed devices were found to be continent at one or more follow-up visits with the low threshold for defining continence possibly contributing to these differing reported outcomes between individual visits.

**Table 3.** Continence rates based on 1-hour pad weight testing, by study visit, n (%)

|  | **4-6 weeks** | **3 months<sup>c</sup>** | **6 months<sup>c</sup>** |
| --- | --- | --- | --- |
| All subjects | 14/23 (60.9%)<sup>a</sup> | 12/21 (57.1%) | 12/21 (57.1%) |
| Subjects with full device extension | 14/15 (87.5%)<sup>a</sup> | 11/14 (78.6%) | 11/14 (78.6%) |
| Subjects with minimal device extension | 0/8 (0.0%) | 1/7 (14.3%) | 1/7 (14.3%) |

Values reported as number of evaluable subjects at each time point (%)
<sup>a</sup> One subject not included in the analysis because they presented with a meatal stenosis at the penis tip due to catheter placement.
<sup>b</sup> Data not yet available for 3 subjects at time of cutoff date for present report.

### Safety

There have been 12 AEs reported to date occurring in 7 (29.2%) subjects in the analysis population for the ARID study (Table 4). This included 1 SAE and 11 AEs, none of which were reported to be related to the study device. There have been no patient deaths. The single SAE and 5 of the AEs were regarded to be definitely related to the radical prostatectomy procedure. The remaining 6 AEs were reported to non-procedure or device related. Overall, 4 of the 12 AEs (33.3%) were considered mild, 7 (58.3%) were considered moderate, and 1 (8.3%) AE was considered severe based on CTCAE V5.0 criteria. The SAE, 3 of the procedure related AEs, and 2 of non-procedure related AEs have been resolved at the time of this report with the remaining AEs still ongoing. There have been no unanticipated adverse device events (UADEs) and all procedure related AEs were consistent with those anticipated with radical prostatectomy.

**Table 4.** Adverse Events.

| Adverse Event | Number of events | Number of subjects (%) | Severity | Procedure Related* |
| --- | --- | --- | --- | --- |
| Meatal stenosis | 1 | 1 (4.2%) | Moderate | No |
| Hypertension | 1 | 1 (4.2%) | Mild | No |
| Respiratory depression/acute respiratory acidosis | 1 | 1 (4.2%) | Severe | Yes |
| Bladder neck lesion | 1 | 1 (4.2%) | Moderate | Yes |
| Post-prostatectomy erectile dysfunction | 2 | 2 (8.2%) | Moderate | Yes |
| Infection | 2 | 2 (8.4%) | Mild | No |
| Urinary infection | 2 | 2 (8.4%) | Mild/<br>Moderate | No |
| Foreign body (plastic clip) found in urinary tract at bladder neck post-procedure | 2 | 2 (8.2%) | Moderate | Yes |

\*none of the adverse events were reported to be device related

The single subject who experienced an SAE desaturated while in recovery after being extubated while also experiencing bradycardia. The subject was put on supplemental oxygen through a face mask and recovered spontaneously without further complications. They were transferred from recovery to semi-intensive care for surveillance and discharged the next day without sequelae. The SAE was deemed to be definitely related to the procedure and not related to the study device.

## Discussion

Post-operative SUI remains a frequent and often debilitating problem for patients undergoing RARP. The ARID study is the first clinical trial to study the use of a novel, absorbable urologic scaffold for the prevention of SUI implanted at the time of RARP with complete device resorption between 6 and 7 months [12]. The present interim analysis of data from this early feasibility study suggests the device is safe and can reduce SUI following RARP. The device was easily inserted and deployed at the time of surgery with no perioperative complications identified.

The observed 6-month results to date from the ARID study compares favorably to other reports evaluating continence following RARP. Pinkhasov et al. [6] reported a 23% continence rate based on a retrospective analysis of 680 patients undergoing RARP and Machioka et al. [13] reported a 52% continence rate at 6 months for their prospective study of 258 subjects undergoing RARP. The outcomes from the present study show improvement compared to this historical data, with a 6-month continence rate of 72.2% for all subjects in this interim analysis of the ARID.

When looking specifically at devices placed optimally, the outcomes of the study improved, both when compared to historical controls and to the data for subjects from the present study when the device was not placed optimally. Optimal device deployment was considered to have occurred when the device was observed to be extended and actively supporting the anastomosed bladder neck. Indeed, when placed optimally (based on video review), 92.9% of patients were continent at 6 months. Optimal implantation of the urologic scaffold requires the device to be fully extended over the anastomosed bladder neck, effectively recreating the prostatic urethra by tubularizing the bladder neck and providing radial support (Fig. 6A). During some procedures, interference from lateral connective tissues which are separated from the bladder during prostatectomy impeded the device’s full extension (Fig. 6B). The manner in which the medial and proximal anchoring sutures were placed for some of the procedures also limited device extension. Additionally, posterior stabilization reconstructions, such as the Rocco Stitch were noted to have interfered with device deployment when excess suture from the reconstruction was incorporated into the anastomosis. All of these sub-optimal conditions minimized device extension, although these were modifiable with simple adjustments in surgical technique (e.g., no Rocco stitch, proper placement of anchoring sutures). While retrospective, it was observed that in nearly all cases the device could have been optimally deployed if the standardized technique were followed. The study demonstrated that correct implantation of the device is important with those subjects in whom the device was fully extended over the anastomosed bladder neck having considerably improved outcomes compared to subjects that had devices that were not optimally extended.

**Fig. 6.**
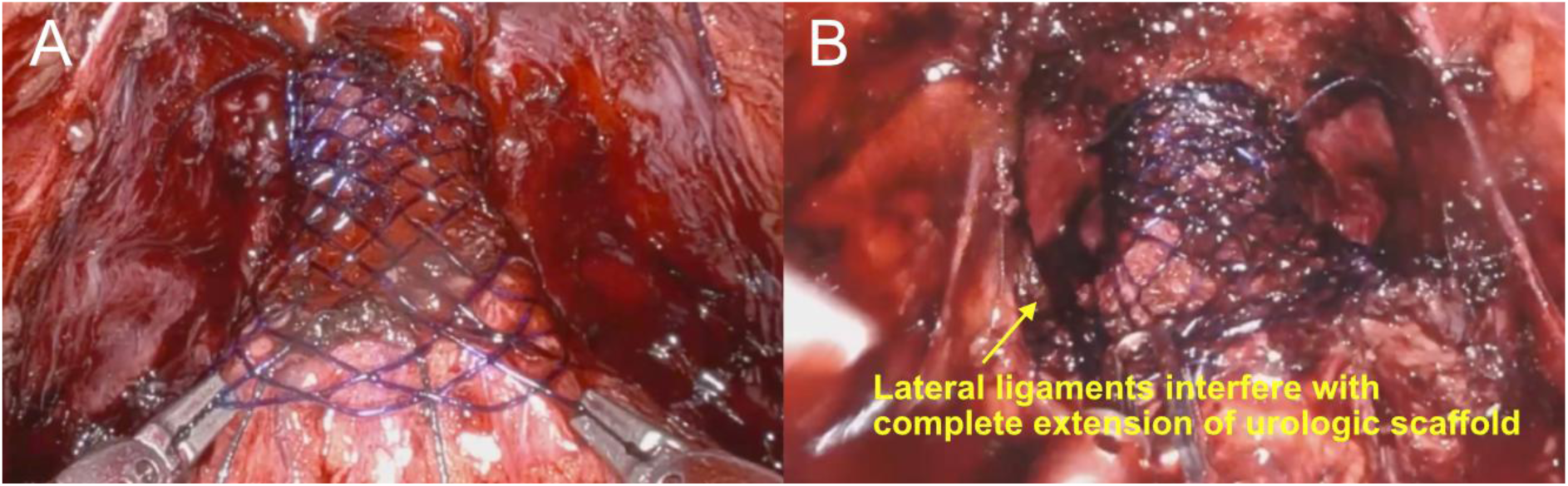
Examples of urologic scaffold deployment. **(A)** optimally deployed scaffold with is fully extended. **(B)** incorrect deployment resutling from interference of the lateral ligaments which prevents full extension of the scaffold.

The lack of early and objective post-prostatectomy pad weight data in the literature makes it challenging to evaluate the comparative benefit the urologic scaffold has on stress incontinence. Pad weight tests are known to have high within-patient variability with Malik et al. reporting that an individual patient’s pad weight values could vary by as much as 51.2 grams on measurements taken on consecutive 24 hour periods [12]. To address this limitation, pad weight results from the ARID study were compared to baseline by establishing a pad weight definition of continence to serve as a responder threshold for both 24-hour and 1-hour pad weight testing, representing the most objective and strictest definition in the literature. The low 5 gram threshold used to define responders for the 1-hour pad weight data possibly made the test sensitive to other sources of moisture that could contribute to measurement error. This is suggested by 5 subjects who had properly placed devices oscillating between responder and non-responder status at differing study timepoints with each achieving continence at one or more study visits. While the use of this 5 gram threshold for determining 1 hour pad weight responder may have resulted in some false negatives, it represents a rigorous definition for continence, and if anything, may be too strict yielding a higher rate of patients determined to be incontinent.

The patient centric outcome of SUI is often understated. Patients suffering from incontinence also experience a decline in quality of life (QoL) and increased regret having undergone the procedure as a result of the emotional, social, occupational and hygienic issues associated with the SUI [14,15]. The effect of urinary incontinence is substantial and requires patients to wear diapers and pads to for most activities of everyday life. This can lead to stigmatization and significant reduction of quality of life including embarrassment, social withdrawal, and physical ailments. Stress urinary incontinence can also impact work productivity due to the need for frequent restroom breaks or a reduction in work hours when the patient experiences severe symptoms. The potential risk of SUI is one of the most feared complications following radical prostatectomy and is often the main driver in patients choosing to not undergo RARP for the treatment pf prostate cancer [16,17]. In addition, the development of stress urinary incontinence following radical prostatectomy has recently been shown to be an independent predictor of increased regret for having undergone the procedure, reduced patient satisfaction, and poorer quality of life [16].

Financial distress or hardship resulting from cancer and its treatment is more frequent among patients who have had increased urinary symptoms following robot-assisted RP [18]. A higher risk of financial toxicity is seen in younger patients, which may be associated with cancer-related reductions in income (e.g., reduced work hours or job loss) that may not occur in older patients who have retirement incomes. This financial toxicity can also lead to treatment regret [19]. For these reasons, developing and deploying an intraoperative mechanism to decrease or prevent SUI in the post-operative period will yield significant patient benefit.

There are several limitations associated with the present study. While subjects were prospectively enrolled, there was the potential for selection bias, and as a result of the lack of a control arm, the ability to compare the results observed in this study to other potential treatment strategies is limited. Additionally, due to the small sample size of this early feasibility study, the inclusion or exclusion of a single subject as a responder in the analysis could have had a proportionally larger impact on the reported continence rate compared to a larger study. The present interim analysis also lacks long-term follow-up data beyond 6 months. These limitations will be addressed by the planned continued reporting of additional results from the ARID study and future reporting of results the ongoing ARID II study (NCT06873581), a large prospective, multicenter, single-blind, randomized controlled study of the urologic scaffold in men undergoing RARP which will follow the subjects for up to two years.

## Conclusion

Novel approaches to prostatectomy are needed to further prevent the development of SUI. Early experience with the absorbable urologic scaffold demonstrates that the device is safe. Based on a rigorous definition of continence used for the present study, the urologic scaffold reduces the rate SUI compared to historical controls. A multicenter, prospective randomized controlled trial using this device which will further define the device’s role in reducing SUI after RARP is currently being conducted.

## Statement of Ethics

The ARID study protocol was reviewed and approved by the Comité de Bioética en Investigación at Pacífica Salud Hospital Punta Pacífica, Panama City, Panama on March 5, 2023. Written informed consent was obtained from the two subjects whose cases are reported in this publication and for the accompanying images.

## Conflict of Interest Statement

MNF and JCG report being paid consultants to Levee Medical and also having equity with the company in the form of stock options. The remaining authors report having no conflicts of interest to declare.

## Funding Sources

Funding for the ARID study (NCT06275945) is being provided by Levee Medical, Inc. who was involved with the design of the study and is also responsible for study monitoring, data collection, the statistical analysis and interpretation of the data.

## Author Contributions

MNF and JCG were involved with the study design and the procedural technique used to place the absorbable urologic scaffold. GE and MY were responsible for coordination of the study. MNF and EB

performed the procedures described by the case reports. MNF drafted the manuscript and coordinated review of the manuscript and manuscript revisions. All authors read and approved the final version of the manuscript.

## Data Availability Statement

The data that supports the findings reported in this paper is not available publicly due to it containing information that could compromise the privacy of study subjects. The data is available from the corresponding author (MNF) upon reasonable request.

## Data Availability

The data that supports the findings reported in this paper is not available publicly due to it containing information that could compromise the privacy of study subjects. The data is available from the corresponding author (J.C.G.) upon reasonable request.

## Notes

### Competing Interest Statement

JCG and MNF report being paid consultants to Levee Medical and also having equity with the company in the form of stock options. The remaining authors report having no conflicts of interest to declare.

### Clinical Trial

Clinical Trial ID: NCT06275945

### Funding Statement

Funding for the ARID study was provided by Levee Medical, Inc. who was involved with the design of the study and is also responsible for study monitoring, data collection, the statistical analysis and interpretation of the data.

### Author Declarations

The ARID study protocol was reviewed and approved by the Comite de Bioetica en Investigacion at Pacifica Salud Hospital Punta Pacifica, Panama City, Panama on March 5, 2023. Written informed consent was obtained from each patient enrolled in the study.

