## Supplementary Material for "Initial Results with an Absorbable Urologic Scaffold to Mitigate Early Urinary Incontinence Following Radical Prostatectomy: The ARID Study"

**Supplementary Table 1.** Summary of Change in 24-Hour Pad Weights Over Time

| Subject Number | Device Placement | Pre-procedure Pad Weight (grams) | Difference in 24 Hour Pad Weight Measurement Post-Radical Prostatectomy vs. Baseline (grams) | | | |
|---|---|---|---|---|---|---|
| | | | Removal | 4-6 Weeks | 3 Months | 6 Months |
| 1 | Full Extension | 15.2 | * | 2.8 | -1.5 | 31.4 |
| 2 | Limited Extension | 10.5 | * | 308.3 | 78.2 | 19.2 |
| 3 | Full Extension | 37.1 | * | -1.7 | -15.5 | -7.7 |
| 4 | Full Extension | 25.7 | * | 16.4 | 96.7 | -3.3 |
| 5 | Full Extension | 19.6 | * | 34.6 | 2.0 | 6.9 |
| 6 | Full Extension | 18.0 | 0.2 | 3.1 | -3.0 | -6.1 |
| 7 | Limited Extension | 37.8 | 434.0 | 815.1 | 619.8 | 419.7 |
| 8 | Full Extension | 23.0 | 3.7 | 3.6 | 0.6 | 0.4 |
| 9 | Full Extension | 37.1 | -27.6 | -12.4 | -11.8 | -16.4 |
| 10 | Limited Extension | 29.3 | 445.1 | 757.6 | 497.7 | 8.7 |
| 11 | Full Extension | 34.2 | 25.0 | 40.3 | 3.8 | 0.2 |
| 12 | Full Extension | 26.8 | 6.4 | 2.0 | -3.2 | -5.3 |
| 13 | Full Extension | 32.2 | 8.6 | -13.7 | 2.0 | 4.2 |
| 14 | Limited Extension | 8.0 | 685.6 | 391.4 | 679.3 | 405.9 |
| 15 | Full Extension | 32.8 | 179.8 | -4.0 | 64.2 | 91.4 |
| 16 | Limited Extension | 9.2 | 232.9 | 428.5 | 285.4 | 195.2 |
| 17 | Full Extension | 28.0 | -1.3 | -2.8 | 0.4 | 2.9 |
| 18 | Full Extension | 22.1 | 17.5 | 62.8** | 27.0 | 31.3 |
| 19 | Limited Extension | 15.5 | 206.0 | 90.6 | 42.6 | 57.6 |
| 20 | Full Extension | 21.1 | -6.3 | -0.3 | -2.5 | -8.3 |
| 21 | Limited Extension | 20.1 | 191.0 | 173.0 | 52.7 | 16.9 |
| 22 | Full Extension | 45.8 | 58.9 | -37.4 | *** | *** |
| 23 | Full Extension | 45.2 | 105.4 | 208.0 | *** | *** |
| 24 | Limited Extension | 2.6 | 320.9 | 319.3 | *** | *** |

* Visit not required for subjects initially enrolled in a previous version of the study protocol
** Not included in the analysis because subject presented with a meatal stenosis at the penis tip due to catheter placement
*** Data not yet available at time of cutoff date for present report

**Supplementary Table 2.** Summary of Change in 1-Hour Pad Weights Over Time

| Subject Number | Device Placement | Pre-procedure Pad Weight (grams) | Difference in 24 Hour Pad Weight Measurement Post-Radical Prostatectomy vs. Baseline (grams) | | |
|---|---|---|---|---|---|
| | | | 4-6 Weeks | 3 Months | 6 Months |
| 1 | Full Extension | 0 | 0.7 | 0 | 0 |
| 2 | Limited Extension | 0 | 41 | 88.7 | 0 |
| 3 | Full Extension | 1.7 | 1.6 | -1 | 9.2 |
| 4 | Full Extension | 1 | 34.8 | 73 | 0.7 |
| 5 | Full Extension | 0 | 0.2 | 0 | 0 |
| 6 | Full Extension | 0 | 2.6 | 10.5 | 0 |
| 7 | Limited Extension | 0.4 | 75.5 | 57.7 | 24.9 |
| 8 | Full Extension | 0 | 0.7 | 0.2 | 0 |
| 9 | Full Extension | 0 | 0.6 | 1.4 | 1 |
| 10 | Limited Extension | 0 | 66.1 | 174.1 | 42.2 |
| 11 | Full Extension | 0 | 2.3 | 0.3 | 2.3 |
| 12 | Full Extension | 0 | 0.3 | 0 | 0 |
| 13 | Full Extension | 0 | 0.7 | 4 | 0 |
| 14 | Limited Extension | 0.1 | 63.4 | 100.8 | 157.5 |
| 15 | Full Extension | 0 | 1.6 | 8.4 | 9.8 |
| 16 | Limited Extension | 0 | 53 | 18.2 | 32 |
| 17 | Full Extension | 16.7 | -15.8 | -16 | -12.9 |
| 18 | Full Extension | 0 | 102.7* | 2.4 | 20 |
| 19 | Limited Extension | 0 | 21.6 | 128 | 72.2 |
| 20 | Full Extension | 0 | 1.7 | 0 | 1.5 |
| 21 | Limited Extension | 0 | 22.5 | 0.4 | 7.2 |
| 22 | Full Extension | 0 | 0 | ** | ** |
| 23 | Full Extension | 0 | 4.8 | ** | ** |
| 24 | Limited Extension | 0.3 | 18.1 | ** | ** |

* Not included in the analysis because subject presented with a meatal stenosis at the penis tip due to catheter placement
** Data not yet available at time of cutoff date for present report
